# Novel Biomarkers of Immune Toxicity from CAR-T Cell Therapy Using Ultrasensitive NULISA™ Proteome Technology

**DOI:** 10.1101/2024.12.18.24319239

**Authors:** Riley Kirkpatrick, Joanne C. Beer, Emily M. Blaum, Agrima Mian, Manishkumar S. Patel, Akansha Jalota, Ishara S. Ariyapala, Qinyu Hao, Wei Feng, Xiao-Jun Ma, Yuling Luo, Brian T. Hill, Neetu Gupta

**Author notes:** Equal contribution (Riley Kirkpatrick, Joanne C. Beer). Correspondence to: Neetu Gupta, Ph.D.; 9500 Euclid Avenue, NE40, Cleveland, OH 44195.

## Abstract

**Translational relevance:** Using serial samples from patients undergoing anti-CD19 CAR T-cell therapy and NULISA^TM^, an assay with attomolar sensitivity, we identify novel plasma proteins associated with severe treatment-associated toxicities. This study not only reveals the evolution of proteomic biomarkers before and during therapy and their association with toxicities, it distinguishes between the induction and recovery phases, and provides insights into factors that may mediate chronic sequelae of CAR T-cell therapy. As the use of CAR T-cell therapy is limited by toxicities, especially in elderly patients or those with pre-existing comorbidities, being able to identify the patients who are at the highest risk for severe toxicities will allow for stratification strategies that would ultimately enable more widespread application of this cutting-edge therapy. Finally, this investigation reveals several potential therapeutic candidates to enable better management and pre-emptive mitigation of acute toxicities.

**Purpose:** The efficacy of anti-CD19 chimeric antigen receptor (CAR) T-cell therapy in large B-cell lymphoma (LBCL) patients is limited by acute toxicities, most notably cytokine release syndrome (CRS) and immune effector cell-associated neurotoxicity syndrome (ICANS). Previous biomarker studies have been constrained by narrow protein panels and limited time points. We employed NULISA^TM^, a novel ultrasensitive assay capable of simultaneously quantifying 204 proteins, to identify temporal proteome associations with acute toxicities in LBCL patients treated with anti-CD19 CAR T-cell therapy.

**Experimental Design:** Plasma samples from 80 LBCL patients who underwent anti-CD19 CAR T-cell therapy, collected before and after cell infusion, were analyzed with NULISA^TM^. Baseline demographics and treatment toxicities, including CRS and ICANS, were graded according to ASTCT consensus criteria. Differential protein abundance, pathway enrichment, and network analysis were performed.

**Results:** Our analysis revealed higher levels of the chemokines CXCL1, CX3CL1, and CCL8 associated with severe toxicity within the first two days of treatment. Thereafter, severe toxicity was marked by more abundant Th2 cell effector cytokines, IL4, IL5 and IL-13, markers of exhaustion, and TNF receptor superfamily proteins CTLA4, PDCD1, CD274, LAG3, TNFRSF1A, TNFRSF1B, and CD40. Finally, patients with severe toxicities showed lower levels of cell growth factors PDGFA and EGF, and the neuronal repair protein BDNF at the resolution stage.

**Conclusions:** This study represents the most comprehensive characterization of proteomic immune response to CAR T-cell therapy to date and identifies novel proteins, pathways, and networks associated with acute toxicity in the initiation and resolution phases, implicating them as potential biomarkers.

## Introduction

Chimeric antigen receptor (CAR) T-cell-based immunotherapy is a novel therapeutic approach against hematologic malignancies in which a patient’s own T cells, isolated and purified from peripheral blood mononuclear cells (PBMCs), are genetically engineered to express CARs that are specific for a target antigen on the tumor. Once a favorable immune environment is created with a brief period of lymphodepletion, the autologous CAR T-cells are infused back into the patient to target and eliminate the tumor (1). CAR T-cell therapy has been used to treat several types of hematological cancers, such as relapsed/refractory large B-cell lymphoma (r/r LBCL), when conventional therapies have proved ineffective (2–4). CAR-T cell therapy is now available for r/r LBCL patients who have disease progression after frontline therapy (5–7). In particular, several anti-CD19 CAR T-cell products including *axicabtagene ciloleucel* (Axi-cel), *tisagenlecleucel* (Tisa-cel), *lisocabtagene maraleucel* (Liso-cel), and *brexucabtagene autocel* (Brexu-cel) have been FDA approved for the treatment of r/r LBCL (2–4,8) or mantle cell lymphoma (9).

While the high response rates associated with CAR T-cell therapy for the treatment of refractory hematologic malignancies are promising, significant limitations and adverse effects prevent its widespread applicability (3). Immune-related adverse events (irAEs) can be severe and possibly life-threatening for the patient, resulting in delay or prevention of a complete return to their pre-treatment functional status (2,10); moreover, irAEs may increase the time of hospitalization and healthcare costs. Because of these toxicities, the potential pool of patients who are eligible to receive CAR T-cell therapy is limited to younger patients without significant comorbidities. The most common irAE following CAR T-cell therapy is cytokine release syndrome (CRS), which occurs in 37% to 93% of patients (11). CRS is most commonly marked by fever, with or without cardiovascular instability, and hypoxia, which can lead to multisystem organ failure in severe cases (12,13). Another common irAE, immune effector cell-associated neurotoxicity syndrome (ICANS), occurs in up to 60% of patients treated with CAR T-cells and can result in disorientation to time or place, impaired attention or short-term memory loss, delirium, headaches, seizures, and cerebral edema with coma (14). Symptoms of CRS can be managed or reversed with intensive monitoring and prompt supportive care, such as administration of tocilizumab, an IL-6–receptor blocking antibody, and/or corticosteroids (15). ICANS is most often treated with steroids, and in severe cases, anti-epileptics or anakinra (IL-1 receptor antagonist) can be used as well (16). The treatment of CRS and ICANS is complex; early, aggressive immunosuppression may limit the effectiveness of CAR-T, yet these irAEs need to be managed to limit resulting morbidity. Together, these long-term toxicities may suggest an ineffective resolution of inflammation or incomplete tissue repair processes. To minimize short-term and long-term morbidity and mortality while preserving the efficacy of this potentially curative therapy, there is a need for identification of toxicity- and resolution-associated biomarkers along the entire timecourse of CAR T-cell therapy.

Laboratory markers of toxicity have been previously identified, including but not limited to C-reactive protein (CRP), ferritin, IL-6, IFN-γ, sIL-2Rα, CXCL8, and CCL2 (6,13,17,18). However, the studies that identified these factors were restricted by relatively narrow panels of target proteins and have often included small patient cohorts and limited time points, making it difficult to appreciate the temporal dynamics of systemic proteome alterations before and after CAR T-cell administration (14,17–19). Here, we employed a novel, ultrasensitive nucleic acid-linked immuno-sandwich assay (NULISA^TM^) technology that can simultaneously quantify 204 inflammation-related proteins in a single sample (20). Plasma samples from 80 r/r patients with large B-cell lymphoma (LBCL), collected at multiple time points before and during treatment with CD19-directed CAR T-cells, were analyzed for associations between the proteome and severity of CRS and ICANS. Using this method, we identified temporally regulated inflammatory or repair proteins, and biological pathways and networks that correlate with severe CRS and/or ICANS.

## Materials and Methods

### Subject recruitment and collection of plasma

The Institutional Review Board of Cleveland Clinic approved the collection of clinical samples in compliance with guidelines for the protection of human subjects as per the Declaration of Helsinki. Written informed consents were provided by all study participants. Peripheral blood samples from 80 r/r LBCL patients were collected at 6 time points before and after CAR T-cell infusion, including the day of apheresis, day of infusion (day 0), and three times post-infusion (day 1-2, day 3-5, day 6-9), and day of discharge (day 11-16). The sample size was set based on sample availability and previous biomarker studies (21,22). Plasma was collected from PBMCs by centrifugation at 1500×g for 30 minutes and stored at −80 °C until subsequent use.

### Patient data

Clinical and demographic information of patients undergoing CAR T-cell therapy were collected by reviewing electronic medical records and maintained in the password-protected and HIPAA-compliant Research Electronic Data Capture (REDCap) system. Gender, age, diagnosis, and prior treatments were recorded. CRS and ICANS grading were recorded on each inpatient day after treatment with CAR T-cell therapy using the ASTCT consensus criteria (23).

### Proteomics analysis

The NULISAseq 200-plex inflammation panel targets mostly inflammation and immune response-related proteins (20). First, plasma samples were centrifuged at 10,000×g for 10 min. Patient sample sets were randomly assigned to one of six 96-well plates. Supernatants from the plasma samples were then analyzed using Alamar’s NULISAseq proteomic platform (details in Supplemental Methods).

### Data processing and quality control

To remove potential technical variation introduced during data collection, data were normalized by dividing NGS read counts in each sample well by that sample’s internal control read counts. Inter-plate normalization was then performed by further dividing each protein target count by the median corresponding target count for the inter-plate controls on that plate. Then, data were rescaled, and log2 transformed to yield NULISA Protein Quantification (NPQ) units for downstream statistical analyses. Quality control (QC) checks indicated one sample with low read counts that was not corrected with normalization; this sample was excluded. All other run and sample QC metrics were within normal ranges.

### Statistical methods

Patients were assigned to binary toxicity response severity groups based on their maximum CRS (maxCRS) grade (2-4 vs 0-1) and maximum ICANS (maxICANS) grade (1-4 vs 0) during the course of the study. The association between maxCRS and maxICANS groups was assessed using Fisher’s exact test. Samples were assigned to time interval bins, including day of apheresis (approximately 21 days prior to the day of CAR T-cell infusion) and days from CAR T-cell infusion, including day 0, day 1-2, day 3-5, day 6-9, and day 11-16. To generate the clustered heatmap, NPQ values for each protein were transformed into z-scores. Proteins and samples within specified time intervals were clustered using agglomerative hierarchical clustering with a Euclidean distance metric and Ward’s minimum variance method. Unless otherwise noted, all statistical analyses were performed using R version 4.3.0. Heatmaps were produced using the ComplexHeatmap package, networks were visualized using igraph and qgraph, and longitudinal protein abundance plots were made using ggplot2 (24).

### Association of longitudinal patterns of protein abundance with CRS or ICANS severity

To assess whether patterns of protein abundance after CAR T-cell therapy varied according to toxicity response severity, linear mixed models were fit for each of the 204 inflammation-related protein targets in the NULISAseq Inflammation Panel. The outcome for each model was protein abundance expressed in NPQ units. Fixed effects included age, sex, categorical time, a binary predictor indicating toxicity response severity (either maxCRS or maxICANS), and the interaction of time with the toxicity severity binary predictor. A random subject intercept was included to account for repeated measures. For each significance test, P-values were adjusted across the 204 proteins using a false discovery rate (FDR) correction, and significance was defined as FDR-adjusted p < 0.05. Significant time-by-toxicity severity interaction coefficients were visualized in a heatmap where protein patterns of differential abundance across time; both toxicity outcomes were clustered using agglomerative hierarchical clustering with a Euclidean distance metric and Ward’s minimum variance method.

### Pathway enrichment and protein interaction network analysis

To determine whether any biological pathways showed significant enrichment, the genes corresponding to significant upregulated and downregulated proteins at each time interval were entered into an enrichment analysis using Metascape (25). The “Multiple Gene Lists” option was selected. The input file included one column for significant upregulated and one for significant downregulated proteins at each time interval. The background column included all 204 proteins in the panel. Physical protein interaction network analysis was performed using STRING (string-db.org) (26). For CRS and ICANS, separate lists for all significant upregulated and downregulated proteins were entered into a network analysis. For protein heterodimers, subunits were entered separately. Network type chosen was physical subnetwork. All other settings were set to the default (e.g., interactions sources included textmining, experiments, and databases; medium confidence level was used).

### Data sharing statement

For original data, please contact the corresponding author

## Results

### Study approach and patient characteristics

Biospecimens and clinical data were collected from 80 patients with r/r hematologic malignancies. Samples were collected at 6 timepoints during the duration of CAR T-cell therapy: day of apheresis (DA), day of infusion (day 0), during therapy duration (day 1-2, day 3-5, day 6-9) and on the day of discharge (day 11-16). Serum samples were then analyzed using the NULISA multiplex assay for inflammation-related proteins (Figure 1A). The median age of the patient cohort was 61 years (range 25-78), and 65% of the patients were male (Figure 1B, Table 1). Figure 1B describes the distributions of maxCRS and maxICANS in our patient cohort. Out of 80 patients, 60 developed CRS of any grade, and 43 developed severe CRS (grade 2-4) during treatment. The average maximum CRS score was 1.34 and the median was 2. Severe ICANS (grade 1-4) occurred in 32 patients. The average maximum ICANS score was 0.75 and the median was 0 (Table 1). Maximum CRS and maximum ICANS groups were significantly associated (Fisher’s exact test p < 0.001), such that patients in the severe group for one toxicity outcome were more likely to be in the severe group for the other outcome (Figure 1B).

**Figure 1.**
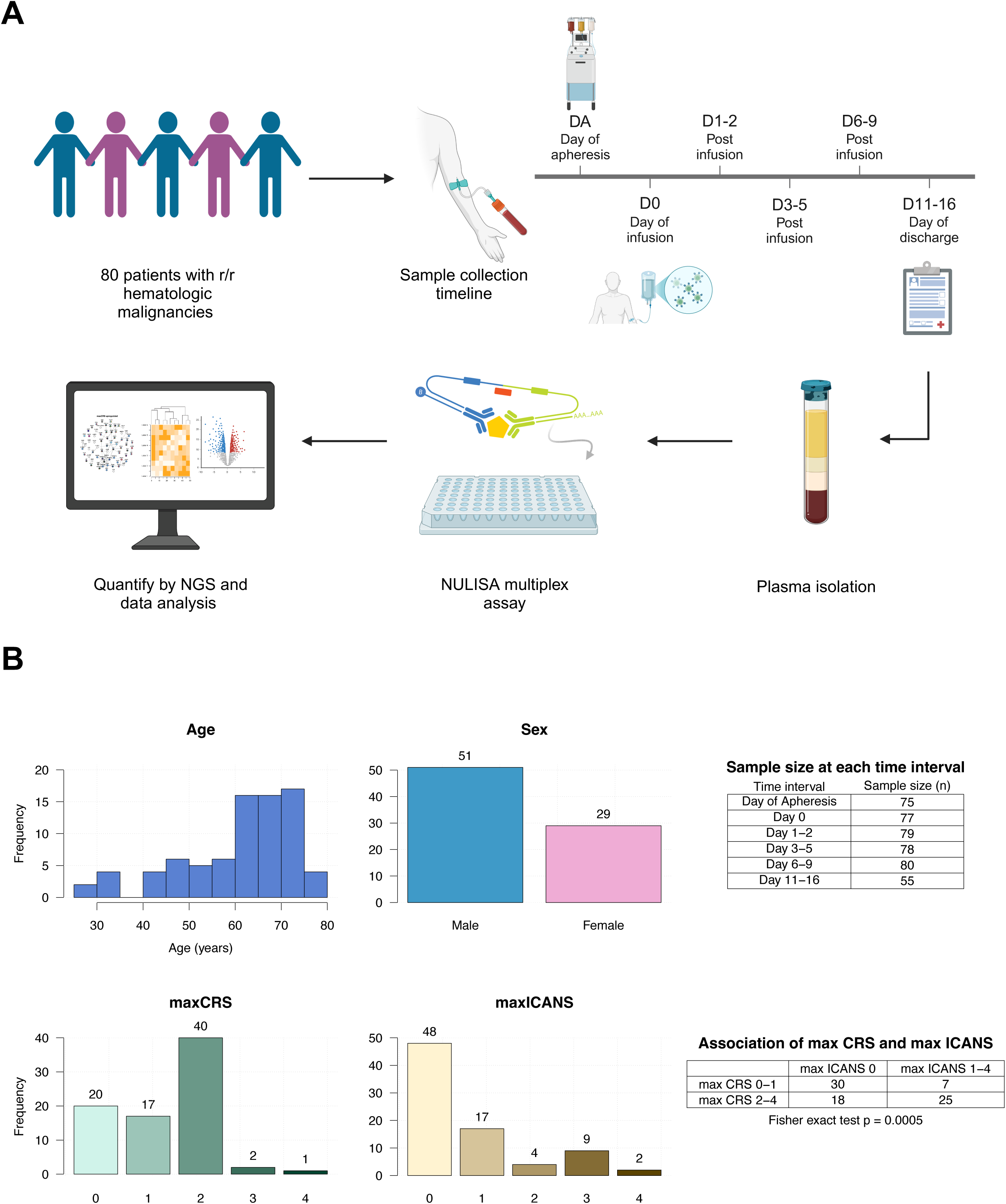
Study schema and sample demographics. **(A)** For each patient, plasma was isolated on the day of apheresis (dA), day of CAR T-cell infusion (day 0), and multiple times post-infusion (days 1-16), plasma proteins were quantified with NULISA, and data subjected to protein abundance, pathway enrichment, network, and association analysis. Diagram was created using BioRender. **(B)** Sample size at each time interval, age, sex, and toxicity outcome distributions. Days are relative to the day of CAR T-cell infusion (Day 0). Day of apheresis occurred approximately 21 days prior to the day of CAR T-cell infusion.

**Table 1:** Baseline and post-treatment clinical characteristics.

|  | **Total n = 80** |
| --- | --- |
| Age (mean (SD), median [min, max]) | 61.1 (12.6), 64 [25, 78] |
| Male (n (%)) | 51 (65%) |
| Disease type (n (%)) |  |
| Diffuse large B-cell lymphoma (DLBCL) | 49 (61.3%) |
| DLBCL transformed from indolent NHL | 19 (23.8%) |
| Primary mediastinal B-cell lymphoma | 6 (7.5%) |
| Mantle cell lymphoma | 3 (3.8%) |
| Follicular lymphoma | 1 (1.3%) |
| Burkitt's lymphoma | 1 (1.3%) |
| Number of prior therapies (median [min, max]) | 3 [2, 7] |
| Prior autologous stem cell transplantation (n (%)) | 54 (67.5%) |
| Type of therapy (n (%)) |  |
| Axicabtagene ciloleucel (Yescarta) | 48 (60.0%) |
| Tisagenlecleucel (Kymirah) | 22 (27.5%) |
| Lisocabtagene maraleucel (Breyanzi) | 8 (10.0%) |
| Brexucabtagene autoleucel (Tecartus) | 2 (2.5%) |
| **Outcomes** |  |
| Maximum CRS<sup>a</sup> (mean (SD), median [min, max]) | 1.34 (0.93), 2 [0, 4] |
| Maximum ICANS<sup>b</sup> (mean (SD), median [min, max]) | 0.75 (1.13), 0 [0, 4] |

<sup>a</sup>Maximum CRS: maximum cytokine release syndrome grade during the study
<sup>b</sup>Maximum ICANS: maximum immune effector cell-associated neurotoxicity syndrome grade
during the study

### Inflammation-related proteins associated with CRS and ICANS severity

Using the NULISAseq proteomic platform, NGS reads were transformed to NULISA Protein Quantification units and subject to downstream analysis (Figure 1A). We first analyzed the longitudinal dynamics of protein abundance prior to and at various times post-CAR T-cell therapy. The clustered heatmap revealed patterns in the abundance of 204 inflammation-related proteins at various times pre- and post-CAR T-cell infusion (Figure 2). Protein clusters were determined using agglomerative hierarchical clustering with an Euclidean distance metric and Ward’s minimum variance method. Here, we refer to each cluster by group number for ease of description. Cluster 1 proteins increased in abundance beginning on day 3-5 and peaked around day 6-9, whereas cluster 2 increased earlier, on day 1-2, and also peaked on day 6-9. Cluster 4 and 5 peaked later, on day 6-9, followed by decreases on day 11-16. Finally, cluster 3 proteins showed some inverted patterns relative to other protein groups, such that patients with high levels in other clusters had lower levels in cluster 3. The heatmap sample annotations suggested an association between high toxicity grades and elevated inflammation markers in protein clusters 1, 2, 4, and 5 and lower levels in cluster 3 (Figure 2).

**Figure 2.**
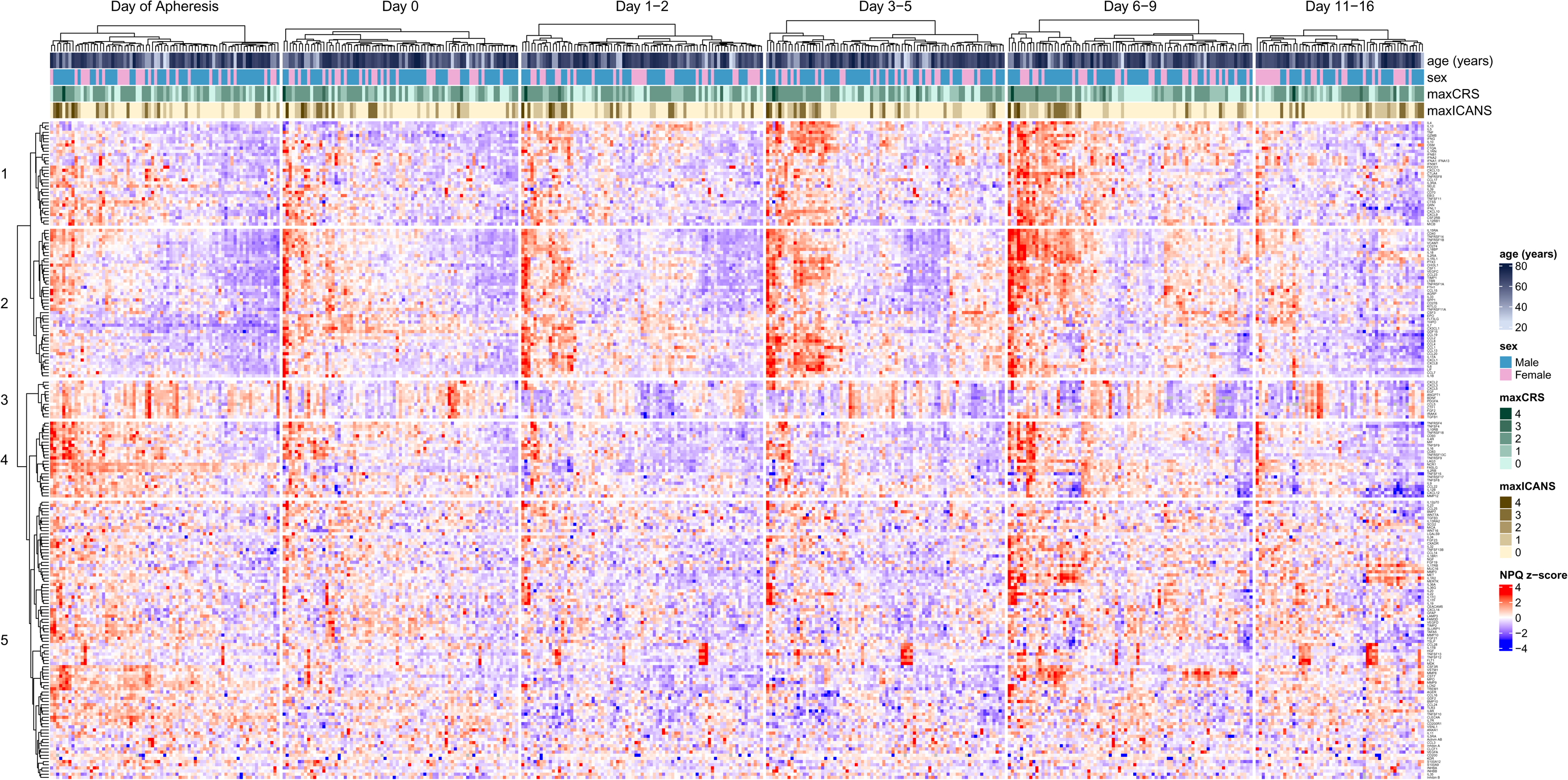
Inflammation-related protein expression. Clustered heatmap shows abundances for 204 proteins measured by NULISAseq Inflammation Panel for samples collected at various time intervals pre- and post-CAR T-cell infusion. Abundances are expressed as standardized NULISA Protein Quantification (NPQ) values. maxICANS: maximum immune effector cell-associated neurotoxicity syndrome grade during the study; maxCRS: maximum cytokine release syndrome grade during the study.

Next, we applied linear mixed effects models to compare the longitudinal proteomic profiles of patients who did or did not develop severe toxicity (maxCRS grades 2-4 or maxICANS grades 1-4). Volcano plots in Figures 3A and 3B and a coefficient heatmap in Figure 4A show upregulated and downregulated proteins that are significantly associated with severe toxicity at different time intervals during CAR T-cell therapy. Overall, of the 204 total proteins in the panel, 71 proteins were upregulated in severe CRS, 45 proteins were upregulated in severe ICANS, and 38 were common to both outcomes. Additionally, 24 proteins were downregulated in severe CRS, 49 proteins were downregulated in severe ICANS, and 13 were common to both outcomes.

**Figure 3:**
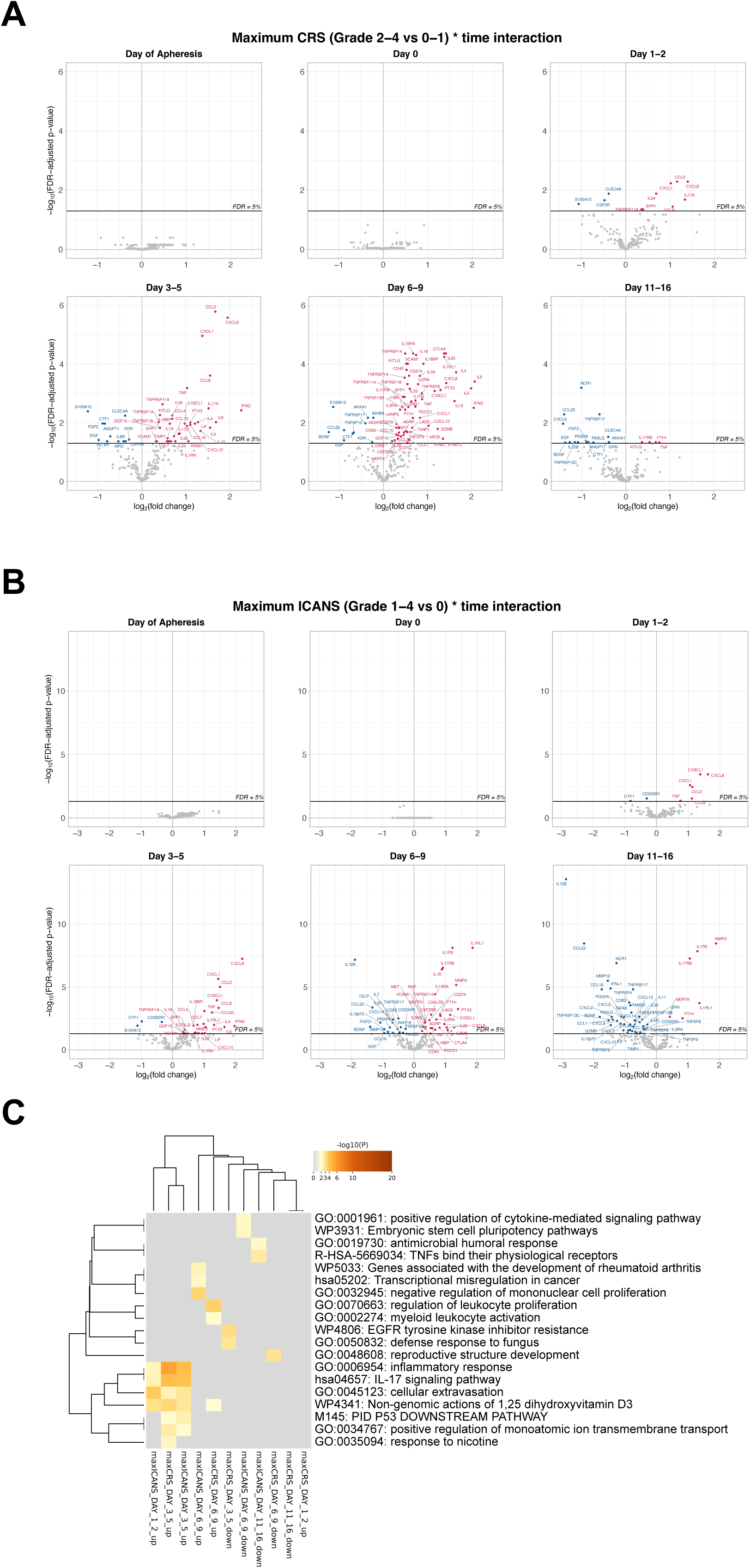
Associations of protein abundances and pathway enrichment with CRS and ICANS after CAR T-cell therapy. Linear mixed models were fit for each of the 204 inflammation-related proteins in the NULISAseq Inflammation Panel. Volcano plots show proteins with significantly different abundances for severe (maxCRS grade 2-4 and maxICANS grade 1-4) versus less severe (maxCRS grade 0-1 and maxICANS grade 0) toxicity for CRS **(A)**, and ICANS **(B)**, at the indicated times during treatment. **(C)** Heatmap showing –log10(p-values) for the top enriched pathways associated with the significant up-or downregulated proteins for maxCRS and maxICANS at the indicated times.

**Figure 4:**
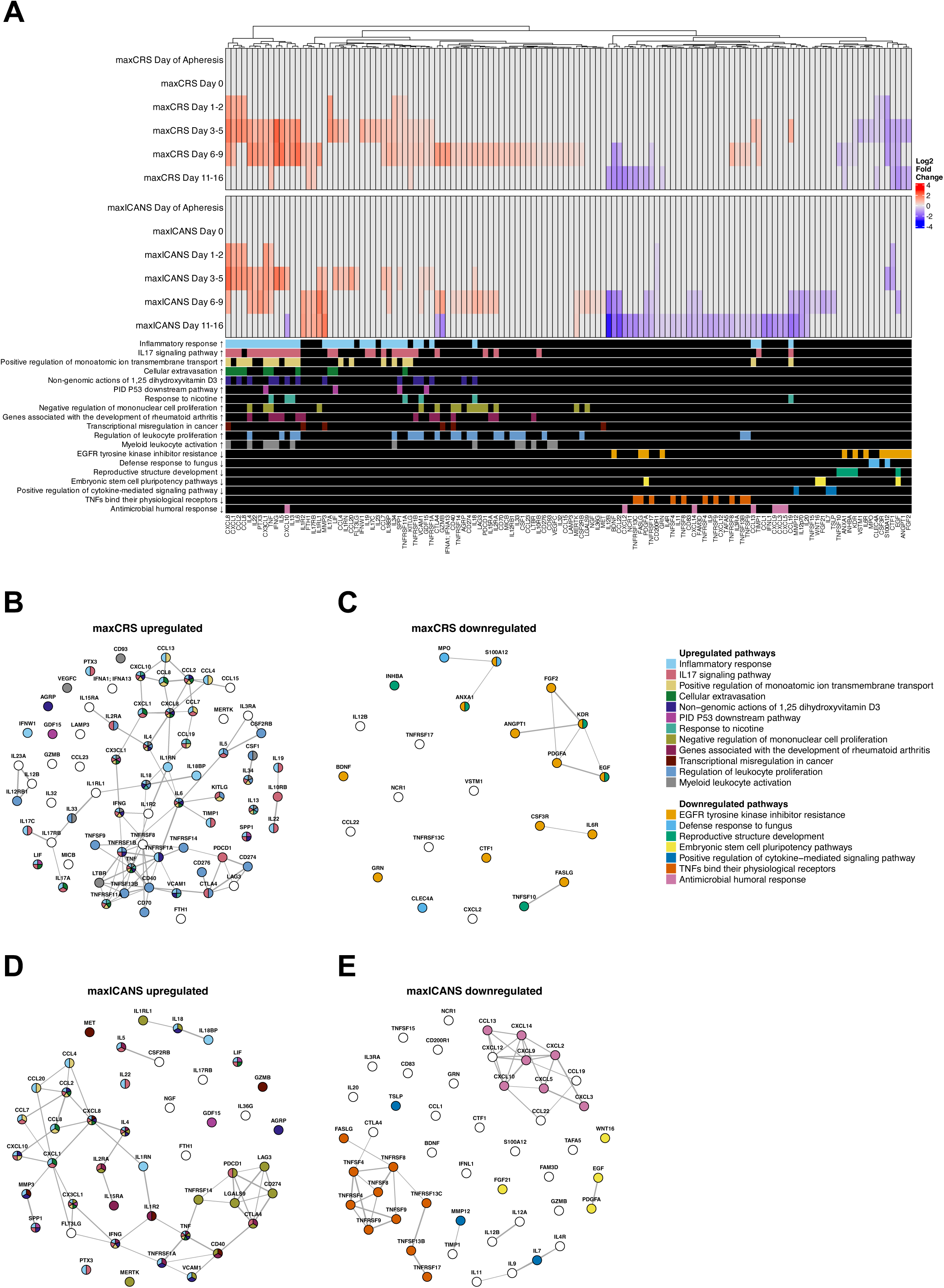
Protein pathways and networks associated with CAR T-cell-induced toxicity. **(A)** Heatmap shows log2 fold change values for all inflammation-related proteins with significant differences by toxicity severity group at various times during CAR T-cell therapy. Annotations at the bottom show which proteins belong to the significantly enriched pathways; arrows indicate up-or downregulation. **(B-E)** Network diagrams show physical protein-protein interactions for significant proteins upregulated in maxCRS **(B)**, downregulated in maxCRS **(C)**, upregulated in maxICANS **(D)**, and downregulated in maxICANS **(E)**. The pathways in which each protein participates are color coded in A-E.

Severe CRS and ICANS revealed some shared patterns in up- and downregulated proteins (Figure 3A, B). Consistent with previous proteomic studies (14,17–19,27), upregulated IL-6, INFG, sIL-2Rα, CXCL8, and CCL2 were correlated with severe CRS and ICANS. These proteins had some of the largest effect sizes, with up to 4-fold increases at peak levels (day 3-5 or 6-9). Additional shared upregulated proteins with large effect sizes included CCL8, CXCL1, and IL5. which peaked on day 3-5 with fold changes above 2.5. S100A12 and CTF1 were downregulated for both outcomes at day 3-5. S100A12, in particular, showed a prolonged downregulation in severe CRS from day 1-2 through day 6-9. On day 6-9, TNFRSF17, BDNF, CCL22, and EGF were downregulated in both severe outcomes, which continued until day 11-16 for TNFRSF17, BDNF, and CCL22, and for EGF as well for severe CRS. Several proteins, including FASLG, TNFRSF13C, and NCR1, were downregulated exclusively at day 11-16 in patients with severe CRS and ICANS (Figure 3A, B). Notably, IL12B and CCL22 were downregulated for both outcomes at day 11-16 and exhibited particularly large changes for severe ICANS, with greater than 4-fold decreases (Figure 3B).

Severe CRS showed unique associations with some of the earliest upregulated proteins, including IL-17A, IL-34, SPP1, and TNFRSF11A, which were elevated within the first two days of therapy (Figure 3A). After this initial period, strongly upregulated proteins for severe CRS included IL-13, IFNA1/IFNA13, and CCL19, which peaked on day 3-5 or 6-9. The earliest downregulated markers for CRS were S100A12, CLEC4A, and CSF3R, showing reductions beginning at day 1-2; CLEC4A remained downregulated through day 11-16, and CSF3R remained downregulated until day 3-5. Unique associations for severe ICANS included early upregulation of CX3CL1 and TNF on day 1-2 (Figure 3B). The earliest downregulated targets on day 1-2 were CD200R1 and CTF1. While showing somewhat fewer upregulated targets on days 3-5 and 6-9 compared to severe CRS, severe ICANS showed more downregulated targets on days 6-9 and 11-16. Severe ICANS also showed a few more strongly upregulated targets at day 11-16, including a nearly 4-fold increase in MMP3. Distinct to severe ICANS were targets with bidirectional effects; in particular, CXCL10 was significantly upregulated at day 3-5, and CTLA4 and GZMB were significantly upregulated at day 6-9; these targets were then significantly downregulated at day 11-16.

### Cellular and molecular pathways associated with severe CRS and ICANS

The genes corresponding to significant upregulated and downregulated proteins at each time interval were entered into an enrichment analysis using Metascape to identify the biological pathways that were significantly associated with severe CRS or ICANS (Supplemental Table 1). Overall, severe CRS and severe ICANS each showed enrichment in nine upregulated pathways, with six pathways in common (Figure 3C, 4A). Three pathways were found to be downregulated in severe CRS, and four in severe ICANS. None of the downregulated pathways were common to the two toxicity outcomes. (Figure 3C). The top enriched pathway associated with both severe CRS and ICANS was the inflammatory response pathway, followed by the IL-17 signaling pathway, which both peaked at day 3-5 (Figure 3C). Other shared upregulated pathways on day 3-5 included positive regulation of monatomic ion transmembrane transport, cellular extravasation, and non-genomic actions of 1,25 dihydroxyvitamin D3 (Figure 3C). These pathways involve several common proteins, including CXCL8, CXCL1, CCL2, CCL8, CX3CL1, IFNγ, and TNFα, which were elevated in patients with severe toxicity by day 1-2 or day 3-5 (Figures 3A, B, 4A).

Severe CRS was associated with uniquely enriched pathways. For example, regulation of leukocyte proliferation was upregulated on day 6-9 (Figure 3C), This pathway includes Th2 cytokines IL-4, IL-5, IL-6, and IL-13 (Figure 4A), which are difficult to detect using traditional methods. Myeloid leukocyte activation was also upregulated at this time interval for severe CRS (Figure 3C, 4A). Downregulated pathways for severe CRS included the EGFR tyrosine kinase inhibitor resistance pathway on day 3-5, involving proteins such as CTF1, EGF, ANGPT1, and FGF2 (Figure 3C, 4A), potentially indicating poor vascular integrity in these patients. Additionally, in severe CRS, defense response to fungus pathway was also downregulated on day 3-5 and is represented by MPO, CLEC4A, and S100A12 (Figure 3C, 4A). Pathways uniquely associated with severe ICANS included upregulation of the negative regulation of mononuclear cell proliferation on day 6-9 (Figure 3C). Furthermore, also unique to ICANS, the TNFs binding to their physiological receptor pathway was downregulated on day 11-16. This manifested as reduced levels of TNF superfamily proteins and receptors (Figure 3B, 3C). Similarly, the antimicrobial humoral response, which includes several chemokines such as CXCL9, CXCL3, and CXCL5, was downregulated on day 11-16 in patients with severe ICANS (Figure 3C, 4A).

### Association of protein networks with severe CRS and ICANS

To identify groups of proteins with previously identified physical interactions, up- and downregulated proteins associated with severe CRS and ICANS were entered into a physical protein association network analysis (Figures 4B-E). In severe CRS, many proteins serving as well-connected hubs (5 or more connections) often belonged to the largest number of enriched pathways. These included CCL2, CXCL8, IL6, TNF, and TNFRSF11A, which were each involved in at least 5 pathways. Two highly interconnected protein groups included a chemokine group involving CCL2, CXCL8, and CCL8, among others, as well as a TNF and TNFR superfamily group (Figure 4B). The largest sub-network for downregulated proteins in severe CRS involved growth factors EGF, PDGFA, FGF2, ANGPT1, and growth factor receptor KDR (Figure 4C), indicating angiogenesis and blood vessel repair as important mechanisms during resolution of inflammation. The protein network upregulated in patients with severe ICANS (Figure 4D) contained fewer targets and was relatively less dense than that upregulated in patients with severe CRS (Figure 4B). The most highly connected (5 or more connections) upregulated proteins in severe ICANS included CCL2, CXCL8, CXCL1, PDCD1, LGALS9, and CTLA4 (Figure 4D). Two notable sub-networks included a chemokine group with CCL2, CCL8, CXCL1, CXCL8, and, importantly, a group of proteins involved in the negative regulation of mononuclear cell proliferation pathway, which included PDCD1, LAG3, LGALS9, CD274, CTLA4, and TNFRSF14 (Figure 4D). Downregulated proteins in severe ICANS showed two strongly connected sub-networks, one consisting largely of TNF and TNFR superfamily proteins and another containing a set of chemokines, many of which were involved in the antimicrobial humoral response pathway, such as CXCL10, CXCL2, CXCL14, CCL13, CXCL9, CXCL3, and CXCL5 (Figure 4E).

### Longitudinal dynamics of protein abundances in the patient cohort

To characterize longitudinal patterns among the proteins associated with severe toxicity, we highlight selected proteins in several important groups throughout our time intervals; these include previously identified markers (Figure 5A), as well as many new markers such as chemokines (Figure 5B), T cell effector cytokines (Figure 5C), exhaustion/feedback regulators (Figure 5D) and mediators of growth/repair (Figure 5E). In addition to CXCL8 and CCL2 (Figure 5A), which have been previously identified, chemokines such as CXCL1, CX3CL1, and CCL8 begin to increase within the first few days of CAR T-cell treatment (Figure 5B). Patients who developed severe CRS and ICANS had higher concentrations of these chemokines within 24 to 48 hours of CAR-T cell infusion compared to those without severe toxicity. Furthermore, the concentrations of T-cell effector proteins, including IL-4, IL-5, IL-13, IL-17A, and IL-18, were higher in patients with severe toxicity on days 3-5, and several of these markers peaked later, at day 6-9 (Figure 5C). Markers of exhaustion such as CTLA4, PDCD1, CD274, and LAG3 and members of the TNF receptor superfamily such as TNFRSF1A, TNFRSF1B, and CD40 (Figure 5D) also peaked at day 6-9. Similarly, these proteins were higher in patients with severe CRS and ICANS during this time interval. Finally, on day 11-16, several mediators of growth, repair, and/or resolution, such as PDGFA, GZMB, BDNF, and EGF, were less abundant in patients with severe toxicity (Figure 5E).

**Figure 5:**
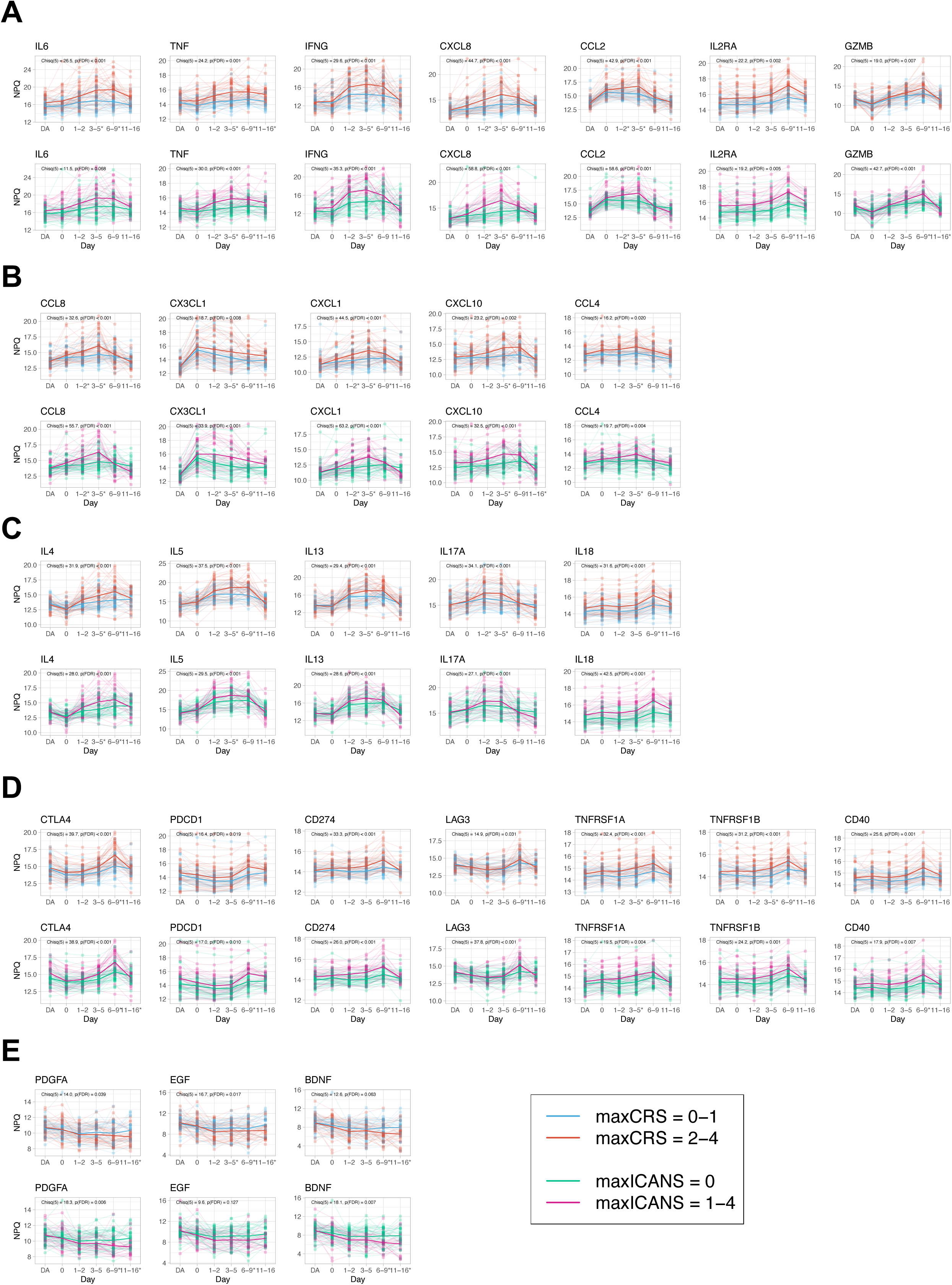
Population-level longitudinal trajectories of protein abundances. Plots show time courses for selected groups of significant proteins, including previously described biomarkers **(A)**, chemokines **(B)**, T cell effector cytokines **(C)**, exhaustion and feedback proteins **(D)**, and growth and repair proteins **(E)**. For each panel, trajectories are colored according to maxCRS (top rows) or maxICANS (bottom rows) toxicity groups. Asterisks next to time intervals on the x-axis denote significant FDR-adjusted differences between toxicity group mean protein abundance at that time point. Chi-square statistics in the upper left corner indicate joint significance of the time-by-toxicity group interaction effect. DA: day of apheresis.

## Discussion

The success and widespread application of CAR T-cell immunotherapy for the treatment of r/r hematological cancers is hindered by life-threatening irAEs, namely CRS and ICANS. While in most cases these complications are reversible, they put patients at risk, require increased personnel for treatment and symptom management, and add additional hospitalization time and healthcare costs. There are several well-known predictors of toxicity, such as high tumor burden, lymphodepletion with cyclophosphamide or fludarabine, and high CAR-T cell dose (28). However, a systematic longitudinal analysis of the pre-treatment, initiation stage and resolution stage proteome and its association with severe toxicities had been missing. In one of the largest cohorts of patients treated with standard-of-care anti-CD19 CAR T-cell product to date, we collected patient samples at multiple time points, including pre-treatment, day of infusion, and during treatment. Using this cohort, we employed NULISA^TM^, a novel ultrasensitive assay that can simultaneously quantify 204 proteins to identify temporal associations of the proteome with severe CRS and ICANS. Our findings not only validate previously identified associations of severe CRS and ICANS with the most commonly studied cytokines IL-6, IFNγ, TNFα, IL-8, IL-10, IL-2Rα, CXCL8, CCL2, MCP-1, GM-CSF, and GZMB (2,13,18,28–35), but reveal several previously undescribed proteins and implicate them in the pathophysiology of inflammation and neurotoxicity related to CAR T-cell therapy.

In the case of both severe CRS and ICANS, shortly after CAR T-cell infusion homeostatic and CAR T-cell-induced chemokines increased, including CX3CL1, which peaked 1-2 days after CAR T-cell infusion, CCL8, CXCL1 and CCL4, which peaked on day 3-5, and CXCL10, which peaked between days 3-9, indicating a temporally organized process of immune cell recruitment that contributes to inflammation throughout the treatment period. By day 11-16, these chemokines returned to pre-infusion levels, indicating a systemic rebound to homeostasis. Similarly, elevated levels of T-cell effector cytokines such as IL-4, IL-5, IL-13, IL-17A, and IL-18 were associated with severe CRS and ICANS and peaked around days 6-9, or earlier for IL-17A, between days 1-5. In particular, the implication of IL-4, IL-5, and IL-13 in the biology of CRS and ICANS is novel due to the difficulty of detecting these cytokines with traditional methods. These Th2 cytokines are involved in humoral immunity, and more specifically, in the repair of tissues after inflammation-related events (36). However, chronic or uncontrolled tissue repair can lead to fibrosis and subsequent organ failure or death, which is characteristic of severe CRS. Interestingly, a recent study reported that during the period of rapid expansion of CAR T-cells (day 6-8 post-infusion) and up to 2 months after infusion, a significant elevation of Th2 cytokines was associated with long-term response to therapy, suggesting the timing of Th2 cytokine upregulation post-infusion is a delicate balance between toxicity and response (37).

At later times, after CAR T-cell infusion, additional pathways were found to be associated with both CRS and ICANS. Interestingly, some mediators of growth and/or repair were downregulated beginning on day 6-9 and extending through day 11-16 in patients with severe toxicity, such as PDGFA, EGF, and BDNF. Among these, BDNF or brain derived neurotrophic factor, is the most prevalent growth factor in the central nervous system (CNS) and plays an important role in neuronal development and plasticity. Here, we identify it as a novel biomarker of CAR T-cell toxicity in both CRS and ICANS. In the CNS, in particular, toxicity to CAR T-cells may be worsened in patients because they do not have access to this vital site-specific growth factor. PDGFA, on the other hand, has been implicated in organ fibrosis and poor vascular integrity (38). Ultimately, these findings suggest that in patients with severe CRS and ICANS, uncontrolled inflammation begins early on post-CAR T-cell infusion, and severe disease progresses or may be prolonged due to insufficient tissue repair processes and/or fibrosis. Insufficient repair and fibrosis, particularly in the CNS, may also explain why patients with more severe ICANS post-CAR T-cell therapy reported worse global cognition after therapy (39). Further, our data implicate T-cell exhaustion in severe CRS and ICANS. Proteins involved in T-cell exhaustion and feedback mechanisms (CTLA4, PDCD1, CD274, and LAG3) peak about a week after infusion (day 6-9) in patients with severe toxicity. This suggests that CAR T-cell exhaustion may not only contribute to their decreased persistence and impaired anti-tumor activity (40) but also to severe toxicity in patients. This finding further underscores the sensitivity of the NULISA^TM^ technology, as markers of exhaustion in circulation have not been identified previously or implicated in the context of CAR T-cell-associated toxicity.

While the pathology of CRS and ICANS are related, our study demonstrates unique proteins that are associated with either severe CRS or ICANS, distinguishing between their underlying biology. In particular, CRS onset typically occurs within a few days of CAR T-cell infusion. The pathology of CRS is thought to be partly due to the expansion of the CAR T-cell population in the peripheral blood and an increase in cytokine levels by the CAR T-cells and bystander immune cells (13). Using network analysis, we were able to identify groups of proteins with previously known physical interactions that were associated with severe CRS. In severe CRS, the largest sub-network for downregulated proteins involved growth factors EGF, PDGFA, FGF2, ANGPT1, and growth factor receptor KDR. Pathway analysis also found that the EGFR tyrosine kinase inhibitor resistance pathway was downregulated at day 3-5; together, these data suggest poor vascular integrity in the pathogenesis of severe CRS. On the other hand, enriched networks for CRS included the TNF and TNFR superfamily group and a chemokine group consisting of CCL2, CXCL8, CLL8 and others, suggesting that interactions between these cytokines, receptors, and chemokines contribute to the inflammatory storm seen in CRS. Typically, the onset of ICANs occurs after CRS and occasionally after CRS has largely resolved, although CRS and ICANS can occur concurrently (31,41). The current understanding of the mechanisms underlying ICANs is incomplete. However, disruption of the blood-brain barrier and cerebral edema caused by cytokine release seem to be significant factors (29,42). Our network analysis of patients with severe ICANS identified two sub-networks that were downregulated – TNF and TNFR superfamily proteins and chemokines involved in the antimicrobial humoral response pathway. In particular, the downregulation of the TNF and TNFR superfamily contrasts with the upregulation of these networks seen in CRS, suggesting that while the two toxicities can be related, there are unique mechanisms at play in the temporal pathogenesis of CRS and ICANS. Upregulated networks in ICANS included proteins that are involved in the negative regulation of mononuclear cell proliferation. Together, these network analyses distinguish between inflammatory markers involved in CRS from those in ICANS.

As CAR-T cell therapy is now being tested as part of frontline treatment for patients with high-risk LBCL, the possibility exists that treatment of this most common type of lymphoma will eventually transition from the application of repeated cycles of traditional anthracycline-based chemotherapy (R-CHOP or similar) to one-time administration of cellular therapy. If CAR T-cell treatment indeed proves to be more effective, the ability to deliver the treatment to the broadest population of patients, including the elderly and/or patients with significant medical comorbidities will continue to be limited by CRS and ICANs. As current management of irAEs is largely in response to symptoms, with IL-6 inhibitors (tocilizumab or siltuximab) used for CRS and glucocorticoids used for ICANS, identification of novel protein mediators of toxicities such as those described here will allow identification of high-risk patient population most likely to benefit from pre-emptive interventions and ultimately enable a broader patient population to receive CAR-T cell therapy with less toxicity, health care cost and more equitable application of this highly innovative therapy. In this regard, our study contributes to a better understanding of the pathophysiology underlying irAEs, which can in turn inform the development of pre-screening measures to identify the most susceptible patients and suggest avenues for prevention and treatment of severe toxicity.

## Data Availability

For original data, please contact the corresponding author

## Acknowledgments

Funding for this study was provided by VeloSano Bike to Cure, Cleveland Clinic Center of Excellence in Lymphoid Malignancies Research and Taussig Cancer Institute. The authors thank patients who consented to participate in this study, as well as clinical nurses and coordinators.

## Authorship Contributions

R.K., A.J. and M.P. collected and processed the blood samples; R.K., J.B., E.M.B., X.M., B.T.H. and N.G. analyzed the data; R.K., J.B., E.M.B. and N.G. wrote the manuscript; B.T.H. and N.G. provided funding support; and N.G. conceptualized and supervised the study.

