## Supplemental Methods and Supplemental Table 1 for "Novel Biomarkers of Immune Toxicity from CAR-T Cell Therapy Using Ultrasensitive NULISA™ Proteome Technology"

^*^ Equal contribution

**Running Title:** Proteomic evolution during CAR T-cell therapy

**Supplemental Methods**

**Proteomics analysis**

For NULISA, the capture antibody (Ab) is conjugated with partially dsDNA containing a poly(A) tail and a target-specific barcode, and the detection Ab is conjugated with another partially dsDNA containing a biotin group and a matching target-specific barcode. When both Abs are incubated with a sample containing the target molecule, an immunocomplex is formed. Immunocomplexes are then captured by paramagnetic oligo(dT) beads and subsequent dT-poly(A) hybridization, and the sample matrix and unbound detection Abs are removed by washing. Immunocomplexes are then released into a low salt buffer. After removing the dT beads, a second set of paramagnetic beads coated with streptavidin is introduced to capture the immunocomplexes a second time. Subsequent washes remove free unbound capture Abs, resulting in essentially pure immunocomplexes on the beads. Then, a ligation mix containing T4 DNA ligase and a specific DNA ligator sequence is added, allowing ligation of the proximal ends of DNA attached to the paired Abs and thus generating a new DNA reporter molecule containing unique target-specific barcodes. The levels of the DNA reporter are then quantified by next-generation sequencing (NGS).

**Supplemental Tables**

**Supplemental Table 1:** Pathway enrichment significant p-values.

**Supplemental Table 1. Pathway enrichment significant p-values**

| **Pathway** | **maxICANS^b^**  **DAY 1-2**  **up** | **maxCRS^a^**  **DAY 3-5**  **up** | **maxICANS^b^**  **DAY 3-5**  **up** | **maxCRS^a^**  **DAY 3-5**  **down** | **maxCRS^a^**  **DAY 6-9**  **up** | **maxICANS^b^**  **DAY 6-9**  **up** | **maxCRS^a^**  **DAY 6-9**  **down** | **maxICANS^b^**  **DAY 6-9**  **down** | **maxICANS^b^**  **DAY 11-16**  **down** |
| --- | --- | --- | --- | --- | --- | --- | --- | --- | --- |
| Positive regulation of cytokine-mediated signaling pathway |  |  |  |  |  |  |  | 5.86E-03 |  |
| Embryonic stem cell pluripotency pathways |  |  |  |  |  |  |  | 4.78E-03 |  |
| antimicrobial humoral response |  |  |  |  |  |  |  |  | 5.95E-03 |
| TNFs bind their physiological receptors |  |  |  |  |  |  |  |  | 1.84E-03 |
| Genes associated with the development of rheumatoid arthritis |  |  |  |  |  | 3.29E-03 |  |  |  |
| Transcriptional misregulation in cancer |  |  |  |  |  | 6.24E-03 |  |  |  |
| Negative regulation of mononuclear cell proliferation |  |  |  |  |  | 3.33E-04 |  |  |  |
| Regulation of leukocyte proliferation |  |  |  |  | 2.38E-04 |  |  |  |  |
| Myeloid leukocyte activation |  |  |  |  | 6.54E-03 |  |  |  |  |
| EGFR tyrosine kinase inhibitor resistance |  |  |  | 6.01E-04 |  |  |  |  |  |
| Defense response to fungus |  |  |  | 1.25E-03 |  |  |  |  |  |
| Reproductive structure development |  |  |  |  |  |  | 9.72E-04 |  |  |
| Inflammatory response | 3.81E-03 | 2.94E-06 | 5.59E-05 |  |  |  |  |  |  |
| IL-17 signaling pathway | 3.61E-03 | 5.43E-05 | 9.08E-05 |  |  |  |  |  |  |
| Cellular extravasation | 1.58E-04 | 3.40E-03 | 1.14E-03 |  |  |  |  |  |  |
| Non-genomic actions of 1,25 dihydroxyvitamin D3 | 8.54E-04 | 3.94E-04 | 1.21E-03 |  | 9.11E-03 |  |  |  |  |
| PID P53 DOWNSTREAM PATHWAY |  | 8.93E-03 | 3.10E-03 |  |  |  |  |  |  |
| Positive regulation of monoatomic ion transmembrane transport |  | 1.95E-03 | 6.58E-03 |  |  |  |  |  |  |
| Response to nicotine |  | 4.84E-03 |  |  |  |  |  |  |  |

^a^Maximum CRS: maximum cytokine release syndrome grade during the study

^b^Maximum ICANS: maximum immune effector cell-associated neurotoxicity syndrome grade during the study
